# Longitudinal co-development of mental and cardio-metabolic health from childhood to young adulthood

**DOI:** 10.1101/2024.10.01.24314697

**Authors:** Serena Defina, Charlotte A.M. Cecil, Janine F. Felix, Esther Walton, Henning Tiemeier

## Abstract

**Objective:** Depressive symptoms and cardio-metabolic risk factors often co-occur. However, our understanding of the potential mechanisms and temporal dynamics underlying their co-development remains elusive.

**Method:** This population-based cohort study examined bidirectional longitudinal associations between depressive symptoms and cardio-metabolic risk factors from age 10 to 25 years, using prospective data from the ALSPAC Study. Participants with at least one (of six) follow-up measurement for each outcome were included in the analyses. We measured depressive symptoms through self- as well as parent-reports, and assessed several cardio-metabolic risk factors (including adiposity measures, lipid profiles and inflammation).

**Results:** Among our 7970 (47% male) participants, we found bidirectional, within-person associations between self-reported depressive symptoms and adiposity (i.e., fat/lean mass index, but not body mass index), across the study period. Adiposity was more stable over time (β [range] = 0.75 [0.54; 0.84]), compared to depressive symptoms (0.26 [0.12; 0.38]), and it had a stronger prospective (i.e., cross-lagged) association with future depressive symptoms (0.07 [0.03, 0.13]) compared to that between depressive symptom and future adiposity (0.04 [0.03, 0.06]). The magnitude of these associations reached its peak between 14 and 16 years. We did not find evidence of cross-lagged associations in either direction between depressive symptoms and waist circumference, insulin, triglycerides, LDL cholesterol or C-reactive protein.

**Conclusions:** These findings suggest a bidirectional relationship between depressive symptoms and cardio-metabolic risk factors, particularly adiposity (i.e., fat/lean mass). Adiposity showed a stronger prospective association with future depressive symptoms, than vice versa, however their relationship revealed more reciprocal than previously thought.

## Introduction

Over 20% of the general population faces at least one depressive episode in their lifetime^1^ and a growing number of adolescents experience depressive symptoms before the age of 20 years.^2,3^ Concurrently, the prevalence of child obesity and related cardio-metabolic risk factors is alarmingly high, affecting one in three children and almost half of young adults.^4,5^ Moreover, depressive symptoms and cardio-metabolic risk factors often co-occur.^1,6,7^ For example, a large meta-analysis showed that people suffering from depression had a 58% higher risk of developing obesity, while individuals with obesity had a 55% elevated risk of developing depression, compared to the general population.^8^

Several potential mechanisms have been proposed to explain this observed comorbidity, both in adults^9,10^ and children^11,12^. However, scientific efforts to empirically model the co-developmental processes that may underlie this comorbidity (i.e., temporal precedence and/or bidirectional relationships) have been sparse, inconsistent, and mostly focused on adult or aging populations^13,14^ and/or genetic liabilities.^15,16^

In the pediatric literature, a small number of studies have investigated longitudinal relationships between body mass index (BMI) and internalizing / emotional problems (an early marker of depressive symptoms).^17-20^ The majority of these studies found higher BMI to precede increases in internalizing symptoms, but not the other way around,^17-19^ in line with some (but not all^15^) Mendelian randomization studies investigating the causal effect of obesity on depression.^16^ A more recent investigation however, did identify reciprocal relationships between BMI and internalizing symptoms, by employing more advanced modeling frameworks, capable of decomposing between- and within-person variances over time.^20^

Importantly, while changes in fat mass are hypothesized to be a key mechanism in these studies, they rely exclusively on BMI measures, which cannot discriminate between fat mass and lean (e.g. muscle) mass, and are thus a suboptimal measure of cardio-metabolic risk.^21-23^ Moreover, existing evidence largely relies on parental reports of depressive symptoms, which may be less sensitive compared to self-reports.^24^ Finally, these studies only investigated relatively short follow-up periods and early developmental windows (i.e., 1 to 5 years, from childhood to early adolescence), leaving the period between adolescence to young adulthood, which is when these conditions typically find their onset, largely unexplored.

To address these gaps, we aimed to characterize the temporal dynamics underlying the (co-)development of depressive symptoms and cardio-metabolic risk as they unfold jointly across 15 years, from the age of 10 to 25 years. We investigated several cardio-metabolic risk factors (including total fat and lean mass) and multi-informant reports on depressive symptoms.

In the spirit of open science, we also provide an open-source interactive web-application *[link redacted]* that can be used, alongside this article, to flexibly explore our results and verify their robustness across multiple outcomes and analytical choices.

## Methods

### Sample and measures

This study is based on data from the Avon Longitudinal Study of Parents and Children (ALSPAC). Pregnant women resident in Avon (UK) with expected delivery dates between 1st April 1991 and 31st December 1992 were invited to take part in the study. The initial number of pregnancies enrolled was 14,541, with 13,988 children who were alive at 1 year of age. When children were approximately 7 years old, additional eligible cases were re-invited, resulting in a total sample of 15,447 pregnancies and 14,901 children who were alive at 1 year of age.^25-27^ Study data were collected and managed using REDCap electronic data capture tools hosted at the University of Bristol.^28^ Please note that the ALSPAC website contains details of all the data that is available through a fully searchable data dictionary and variable search tool (http://www.bristol.ac.uk/alspac/researchers/our-data/).

Ethical approval for the study was obtained from the ALSPAC Ethics and Law Committee and the Local Research Ethics Committees.

#### Depressive symptoms

Depressive symptoms were repeatedly measured using the Short Mood and Feelings Questionnaire (SMFQ)^29,30^. The instrument includes 13 items referred to the past two weeks and scored between 0–2 (i.e., “not true” / “sometimes true” / “true”). A summary score ranging between 0–26 was computed at each occasion, with higher scores indicating greater depressive symptoms (*Figure 1A*). The SMFQ was administered to the child/young person on six occasions between the ages of 10 and 25 years (at the median ages of 10.6, 12.8, 13.8, 16.6, 17.8, and 23.8 years). The questionnaire was also completed by participants’ parents (most commonly mothers) on four additional occasions (when children were on average 9.6, 11.7, 31.1, and 16.7 years old) which were used in secondary analyses.

**Figure 1.**
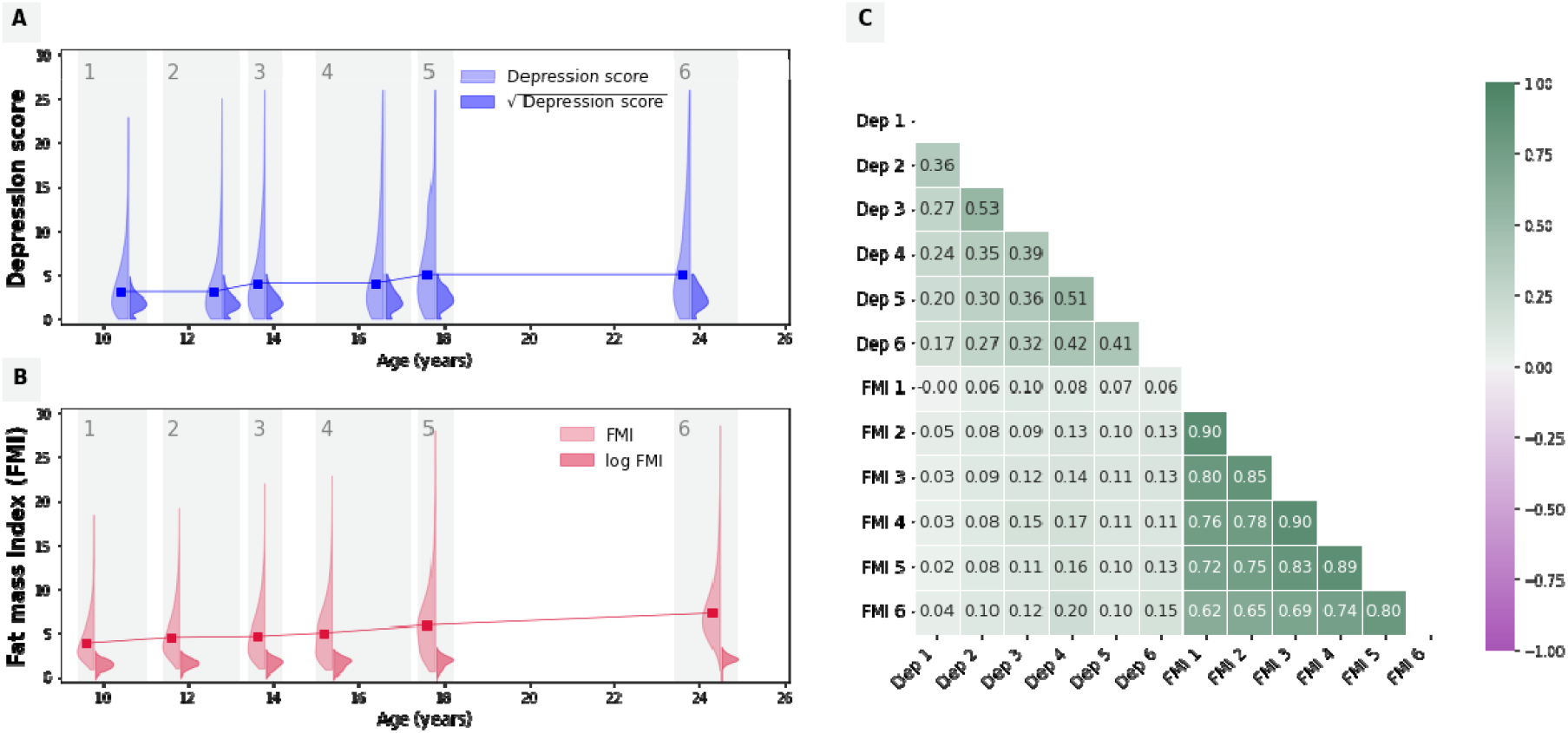
Main outcomes descriptives. **(A, B)** The distribution of observed values for SMFQ depressive scores (A) and fat mass index (B) is presented on the y-axis against measurement time (x-axis). In the violin plots, lighter colors are used to represent the original value distributions, while darker colors represent the same variable distributions and after data transformation was applied (i.e., square root for depressive symptoms scores and log transformation for FMI). The line graph connects the median points (in the original data scale) at each timepoint. **(C)** The univariate, pairwise Pearson correlation coefficients between each repeated measure of depressive symptoms (Dep) and fat mass index (FMI).

#### Cardio-metabolic risk markers

The primary cardio-metabolic measure examined in this study was fat mass index (FMI; *Figure 1B*), computed as participants’ total body fat mass divided by their squared height (kg/m^2^). Total body fat mass was derived from whole body dual energy X-ray absorptiometry (DXA) scans at six occasions (at median ages of 9.8, 11.8, 13.8, 15.4, 17.8, and 24.5 years).^31^

Ten other cardio-metabolic risk factors were further examined in secondary analyses, including lean mass index (LMI), body mass index (BMI), waist circumference, android fat mass, high-density lipoprotein (HDL) and low-density lipoprotein (LDL) cholesterol levels, triglycerides, insulin, and C-reactive protein (CRP) (see *eMethods1, eFigure1*).

#### Sample demographics

Participant sex was measured at birth. Participants’ parents further reported on their ethnical identity (at recruitment) and their educational attainment (when children were 5 years old).

### Statistical analyses

Analyses were conducted using R version 4.2.2^32^; model specification and fit was implemented in lavaan (version 0.6–16)^33^. All scripts are publicly available *[link redacted]*.

#### Data pre-processing

To ensure optimal model conversion, we first performed data cleaning by setting extreme outlier values (i.e., > 5 interquartile ranges above the third quartile or below the first quartile) to missing. We then transformed the data to reduce skewness, using a square root transformation (for depressive symptoms scores; *Figure 1A*) or a ln transformation (for cardio-metabolic risk markers; *Figure 1B*). Finally, we performed min-max normalization (i.e., subtracting the minimum value from each observation, and dividing this difference by the value range) to rescale of all variables to a [0, 1] range.

#### Main analyses

We fit a lag-1 random-intercept cross-lag panel model (RI-CLPM)^34^ to characterize the relationship between self-reported depressive symptoms and fat mass index across 6 timepoints (∼10, 12, 14, 16, 18 and 24 years). The model was specified as a structural equation model composed of four parts (see also *eMethods2, eFigure2*):

1. A *between-person* part, consisting of the “random intercepts” (η_*dep*_ and η_*FMI*_). These are latent variables that have each measurement occasion as indicator and factor loadings set to 1. They reflect stable (i.e., “time-invariant”) between-person differences (e.g., some children may have systematically higher fat mass over time compared to others).
2. A *within-person* part, consisting of “within-unit fluctuations”: time-specific residual terms specified as latent variables with factor loading set to 1, and (measurement error) variances set to 0. They represent random changes that make observations unique, allowing individuals to differ (from themselves) at each occasion. For example, these could reflect a life event that raises/lowers a person’s depression at a given time *t*.
3. The *(lag-1) regressions* between the within-unit components: i.e. the auto-regressive and cross-lagged terms.
  a. auto-regressive terms quantify the persistence (or “inertia”) of a construct, i.e. its tendency to retain its state over time. For example, AR_dep_ captures the proportion of past depression that persists directly to the next measurement occasion.
  b. In contrast, cross-lagged relations measure the proportion of past variance in one variable that is reflected in the *other* variable at the next measurement occasion, and are therefore used to infer (Granger) causality. For example, CL_dep_ indicates how much within-person variance in depression at time *t* is uniquely explained by FMI at timepoint *t –1* (controlling for the persistence of past values of depression).
4. *Covariances* in the between- and within-person part.
  a. To control for between-person trends that may confound the (within-person) system dynamics reflected by auto-regressive and cross-lagged terms, the covariance between η_*dep*_ and η_*FMI*_ is freely estimated.
  b. Similarly, because within-unit fluctuations may be non-independent (e.g., when a random change affects both variables simultaneously) this is modelled by estimating their covariance within each wave.

Full information maximum likelihood (FIML) estimation was used to account for missing patterns that may not conform to MCAR. Coefficients were standardized and conventional robust standard errors were used to compute 95% confidence intervals (95%CI).

Model fit was evaluated using the Root Mean Square Error of Approximation (RMSEA), the Comparative Fit Index (CFI), the Tucker Lewis Index (TLI), and the Standardized Root Mean Square Residual (SRMR). We considered model fit to be adequate when: RMSEA ≤ 0.05, CFI and TLI ≥ 0.95, and SRMR < 0.08.^35^

### Secondary analyses

We further conducted two sets of secondary analyses.

a. We replaced FMI with each of 10 alternative cardio-metabolic risk markers, including measures of adiposity (i.e., BMI, LMI, android fat mass and waist circumference), lipid profiles (i.e., HDL and LDL cholesterol levels, and triglycerides), insulin levels and inflammation (i.e., CRP).
b. We replaced self-reported depressive symptoms scores with maternal reports (at the available timepoints). Note that the project web-application *[link redacted]* offers the opportunity for researchers to interact with model settings and examine the robustness of findings against violations of modelling assumptions, such as the temporal stability of the between- and within-person components (*eMethods2*).

## Results

### Descriptive statistics

Sample descriptives are presented in *Table 1* and *Figure 1*. The main analytical sample consisted of 7970 (47% male) participants, who had at least one measurement of depressive symptoms and FMI.

**Table 1.** Sample descriptives.

|  | **Total sample**<br>**(n = 7970)** | **Male participants**<br>**(n = 3769)** | **Female participants**<br>**(n = 4185)** |
| --- | --- | --- | --- |
| **SMFQ depressive symptom score**, median (range) [% missing values] |  |  |  |
| 10 years | 3 (0-23) [15%] | 3 (0-23) [14%] | 3 (0-21) [17%] |
| 12 years | 3 (0-25) [19%] | 3 (0-25) [17%] | 3 (0-24) [21%] |
| 14 years | 4 (0-26) [24%] | 3 (0-26) [21%] | 4 (0-26) [26%] |
| 16 years | 4 (0-26) [43%] | 3 (0-26) [51%] | 5 (0-26) [36%] |
| 18 years | 5 (0-26) [45%] | 4 (0-26) [51%] | 6 (0-26) [40%] |
| 24 years | 5 (0-26) [54%] | 4 (0-26) [66%] | 5 (0-26) [42%] |
| **Fat mass index**, median (range) [% missing values] |  |  |  |
| 10 years | 3.8 (0.7-18.4) [50%] | 3.0 (0.7-15.4) [49%] | 4.4 (1.0-18.4) [51%] |
| 12 years | 4.4 (0.9-19.1) [51%] | 3.8 (0.9-16.8) [50%] | 5.0 (1.1-19.1) [51%] |
| 14 years | 4.6 (0.8-21.9) [24%] | 3.1 (0.8-17.7) [22%] | 5.7 (1.5-21.9) [27%] |
| 16 years | 5.0 (0.7-22.8) [36%] | 2.8 (0.7-20.0) [36%] | 6.4 (1.5-22.8) [36%] |
| 18 years | 5.8 (0.5-27.9) [40%] | 3.4 (0.5-24.1) [44%] | 7.1 (0.5-27.9) [36%] |
| 24 years | 7.2 (0.6-28.5) [53%] | 5.7 (1.8-23.2) [63%] | 8.0 (0.6-28.5) [44%] |
| **Sex**, n (%) |  |  |  |
| Male | 3769 (47%) |  |  |
| Female | 4185 (53%) |  |  |
| **Ethnic background**, n (%) <sup>a</sup> |  |  |  |
| Non-white | 280 (4%) | 134 (4%) | 146 (4%) |
| White | 6718 (96%) | 3215 (96%) | 3503 (96%) |
| **Maternal education**, n (%) <sup>b</sup> |  |  |  |
| No education | 210 (3%) | 89 (3%) | 121 (4%) |
| Medium | 4836 (78%) | 2374 (78%) | 2462 (77%) |
| High | 1164 (19%) | 561 (19%) | 603 (19%) |
| **Paternal education**, n (%) <sup>b</sup> |  |  |  |
| No education | 349 (7%) | 165 (6%) | 184 (7%) |
| Medium | 3475 (66%) | 1676 (65%) | 1799 (66%) |
| High | 1469 (28%) | 730 (28%) | 739 (27%) |

<sup>a</sup> Ethnic background: "White" if both parents identified as "White"; "Non-white" if either parent identified as "Black Caribbean", "Black African", "Other black", "Indian", "Pakistani", "Bangladeshi", "Chinese", or "Other".
<sup>b</sup> Maternal/Paternal education: medium = "CSE", "Vocational", "O level", "A level"; high = "(College or university) degree". Categorization based on ISCED 2011.

Both depressive symptoms and FMI increased slightly with age (Figure 1A-B); their cross-sectional correlations ranged from 0.00 to 0.17 (r mean = 0.10; Figure 1C).

Girls had systematically higher FMI compared to boys across timepoints, and they reported higher depressive symptoms scores from the ages of 14 years onwards.

### Main results

Results of the main analyses, examining the co-development of depressive symptoms and FMI, are summarized in *Table 2* and *Figure 2*. The model showed good fit (χ^2^(37) = 356.77, *p*<0.001; RMSEA [95%CI]□=□0.033 [0.030-0.036]; CFI = 0.991; TLI = 0.983; SRMR = 0.027).

**Table 2.**
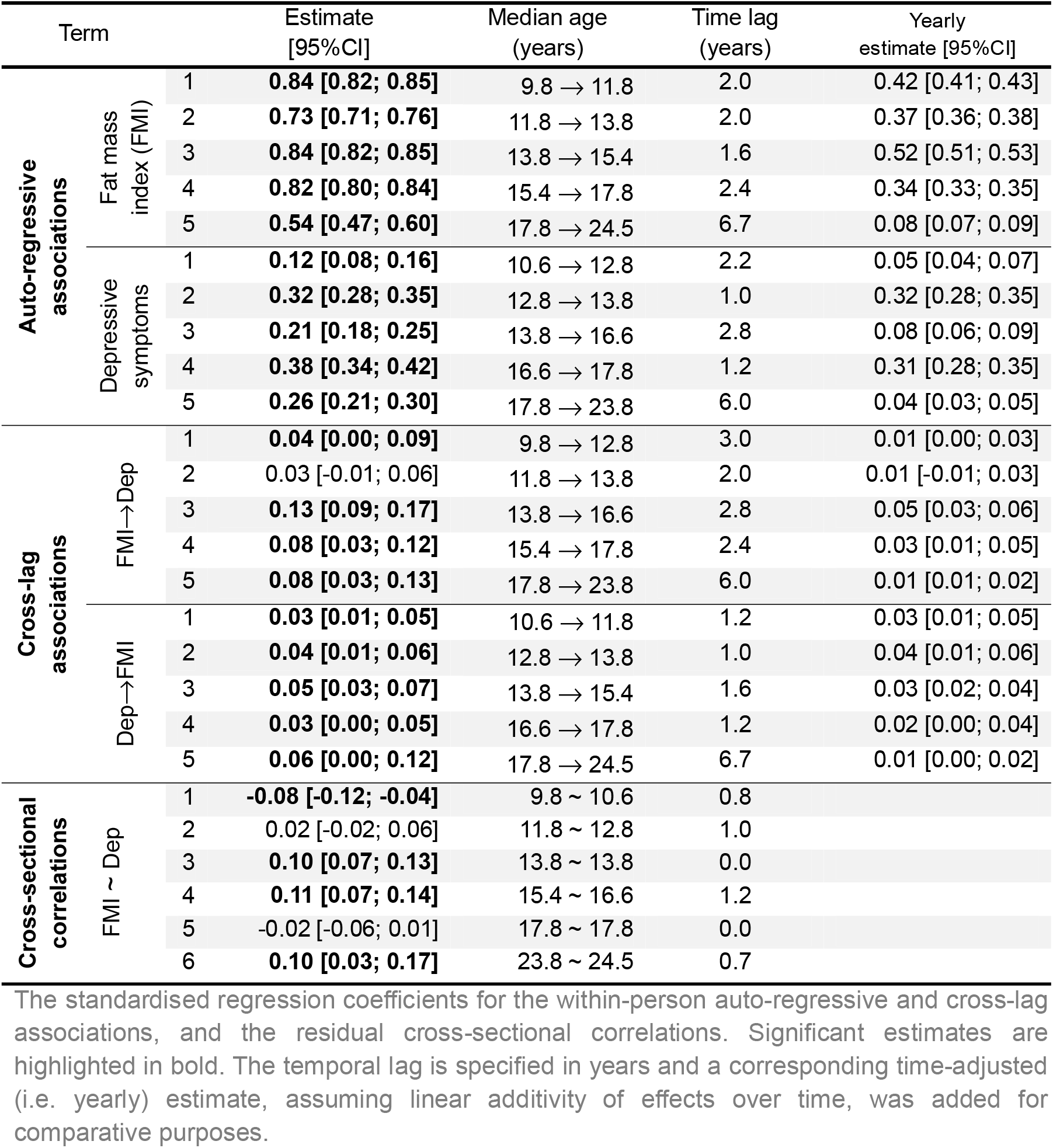
Auto-regressive and cross-lag associations between depressive symptoms and FMI.

**Figure 2.**
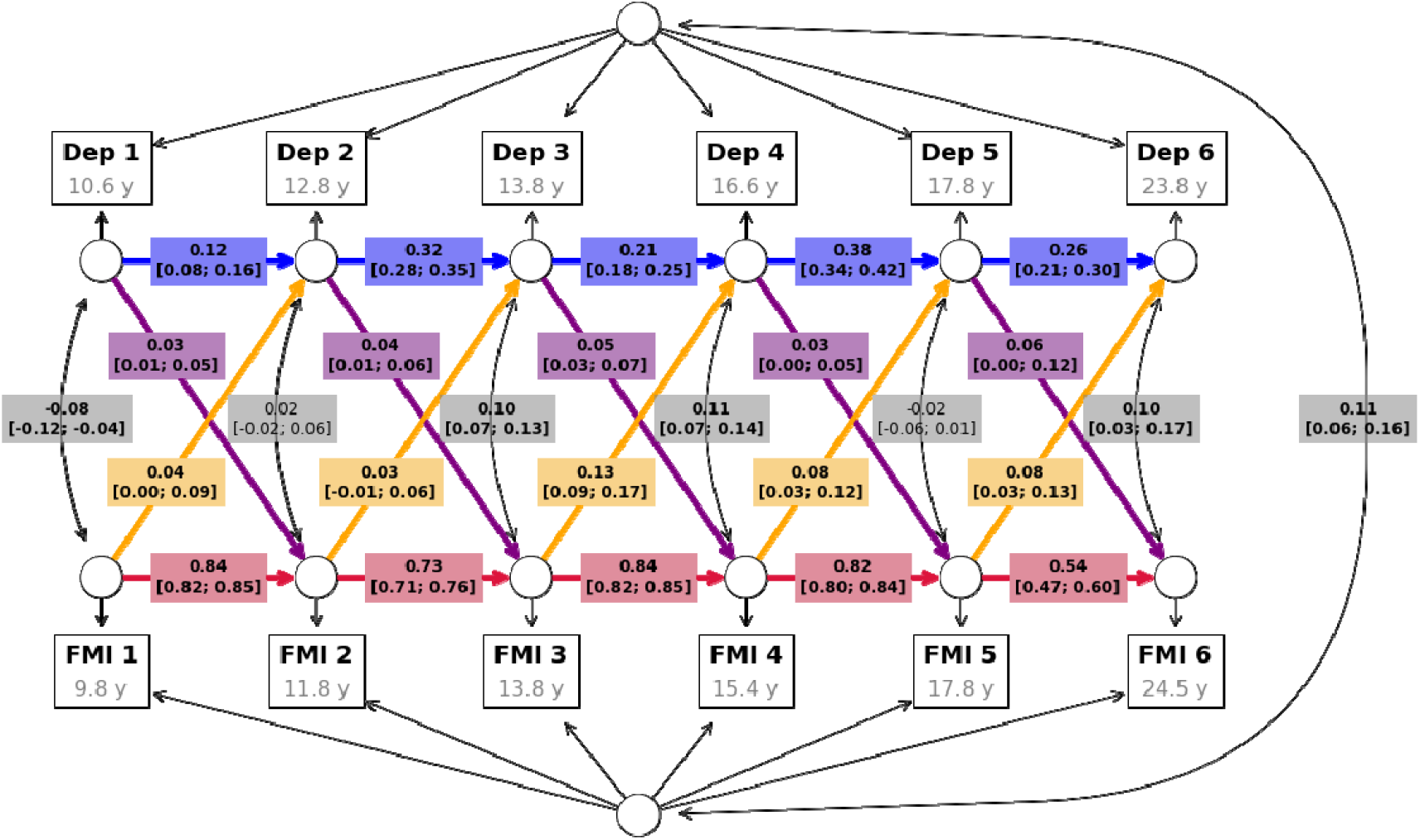
RI-CLPM of depressive symptoms and FMI. Results of the main analysis (i.e., beta values and their 95% confidence intervals) are displayed within their implied directed graph. Autoregressive associations are shown in red for FMI and in blue for depressive symptoms (Dep), while cross-lag effects are shown in yellow for FMI to depressive symptoms paths and in purple for depressive symptoms to FMI paths. Grey boxed enclose the correlation coefficients (and their 95% confidence intervals) between the two random intercepts, as well as the cross-sectional correlations between the two outcomes at each measurement wave.

**Figure 3.**
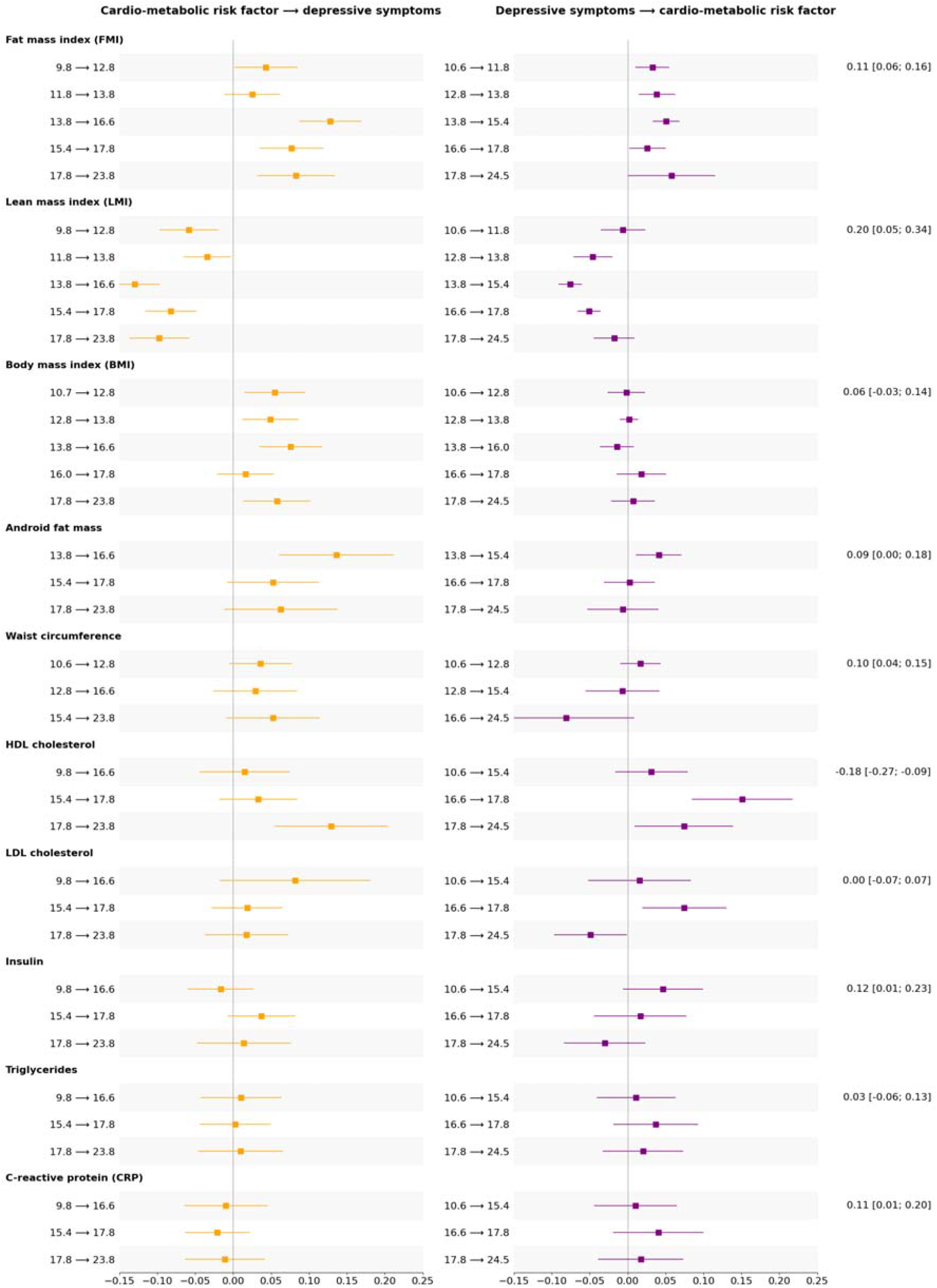
Cross-lag associations between depressive symptoms and cardio-metabolic risk factors. The standardised regression coefficients for the within-person cross-lag associations (and their 95% confidence intervals) are presented along the x-axis, for each alternative cardio-metabolic risk factor, listed on the y-axis. The temporal lag each estimate refers to is specified in years on the y-axis. Association estimates from cardio-metabolic risk factor to lagged depressive symptoms are presented in yellow on the left and those between depressive symptoms and lagged cardio-metabolic risk factor are shown in purple on the right. In the last column on the right of the graphs, the estimated correlation coefficient between the random intercepts of each construct (and its 95% confidence interval) is reported.

We found positive auto-regressive associations, indicating substantial within-person stability over time in FMI (mean β [range] = 0.75 [0.54; 0.84], SE = 0.016) and, to a lower extent, in depressive symptoms (β [range] = 0.26 [0.12; 0.38], SE = 0.022).

After accounting for within-person (i.e., autoregressive associations) and between-person stability (i.e., random intercepts), the following within-person cross-lag dynamics emerged from our models (Table 2): higher FMI was associated with increased subsequent depressive symptoms across the study period, except between 12 and 14 years (β [range] = 0.07 [0.03, 0.13], SE = 0.030); higher depressive symptoms were associated with increased subsequent FMI, although these associations were weaker on average, compared to those between FMI and future depressive symptoms (β [range] = 0.04 [0.03, 0.06], SE = 0.01).

We additionally found a positive correlation between the random intercepts (r□[95%CI] =□0.11 [0.06-0.16]), suggesting some stability of between-person associations between depressive symptoms and FMI.

### Secondary analysis results

Please visit the project web-application *[link redacted]* for an interactive report of all results obtained from secondary analyses. We highlight and summarize below a few key findings.

#### Other cardio-metabolic risk factors

The pattern of reciprocal cross-lag paths identified in the main analysis was remarkably consistent when lean mass (rather than fat mass) index was examined as a cardio-metabolic risk factor (average standardized βs for LMI to depressive symptoms = -0.08, SE = 0.029; depressive symptoms to LMI = -0.04, SE = 0.007). When BMI was included in the model instead, only the prospective association between BMI and depressive symptoms remained (β = 0.05, SE = 0.035), while depressive symptoms did not seem to affect future BMI (β < 0.01, SE = 0.008). Android fat mass and depressive symptoms showed reciprocal associations only between 14 and 16 years (android fat to depressive symptoms = 0.14 [0.06; 0.21]; depressive symptoms to android fat = 0.04 [0.01; 0.07]).

Surprisingly, a positive within-person reciprocal association between depressive symptoms and HDL (but not LDL) cholesterol levels was detected, between 16 and 25 years (see *Table 3*). In contrast, the correlation between the random intercepts of depressive symptoms and HDL cholesterol was negative (−0.18 [-0.27; -0.09]).

We did not find evidence of association in either direction when waist circumference, insulin, triglycerides, or CRP were considered in relation to self-reported depressive symptoms.

#### Maternal reports of depressive symptoms

The reported findings were relatively consistent (albeit weaker) when maternal reports of depressive symptoms were used in place of self-reports (average standardized βs for FMI to depressive symptoms = 0.09, SE = 0.041; depressive symptoms to FMI = 0.04, SE = 0.013; see eFigure 3). Please visit the project web-application *[link redacted]* for a complete report of these findings.

## Discussion

In this population-based study, we characterized the between- and within-person associations governing the co-development of depressive symptoms and cardio-metabolic risk factors, from age 10 to 25 years. Specifically, we found bidirectional, within-person associations between depressive symptoms and adiposity (i.e., fat/lean mass index, but not body mass index). Adiposity was more stable over time, compared to depressive symptoms, and it had a stronger prospective association with future depressive symptoms compared to that between depressive symptom and future adiposity.

### The co-development of depressive symptoms and adiposity: fat vs. weight measures

Interestingly, the pattern of reciprocal associations between depressive symptoms and adiposity was only evident when using fat and lean mass measures, not BMI. The prospective relationship between earlier depressive symptoms and later BMI was not detectable, similar to what has been reported in previous genetic^16^ and longitudinal epidemiological studies conducted in children.^17-19^

This is important because, despite its popularity, BMI is not the best indicator of cardio-metabolic risk (e.g., a high BMI may also reflect high muscle mass, and a normal BMI does not exclude a high body fat percentage).^21^ Compared to fat-based measures, BMI was also a weaker predictor of depressive symptoms (as well as other physical heath complications) in older adults.^9,36-38^

Based on this evidence, we recommend the use of fat or lean mass measures when assessing the relationship between adiposity and depressive symptoms, as relying on BMI only may lead the erroneous conclusion that increased adiposity is only an antecedent (rather than a consequence) of early-onset depressive symptoms.

### Maternal-vs. self-reports of depressive symptoms

We found a generally similar pattern of relationships between depressive symptoms and adiposity, when using maternal reports of depressive symptoms (rather than self-reports). However, associations were weaker when using maternal reports and they were less consistent across timepoints. Previous studies have relied solely on maternal reports when investigating relationships with child adiposity, which may have further contributed to inconsistencies in the reported timing, direction, and magnitude of these associations.

### The wheels of a “vicious cycle”

Several potential mechanisms could modulate and/or explain the bidirectional within-person dynamics highlighted in this study. For example, higher adiposity may affect psychological well-being, through reduced self-esteem, and increased feelings of shame, isolation, and body image dissatisfaction,^39^ especially during adolescence. Adipose tissue is also an endocrine organ, involved in the production of oestrogen and pro-inflammatory cytokines^40^, which could induce symptoms of depression such as anhedonia, fatigue, sleep problems and social withdrawal.^41^ In turn, depressive symptoms may increase the risk for subsequent adiposity through poorer diet^42^, sleep quality and sedentary behavior.^43^

Interestingly, in addition to these within-person associations (which are directional and time-specific) we also found some stable between-person associations between depressive symptoms and adiposity. This is an indication that a consistent portion of the comorbidity burden can be better explained by “shared risk factors” that are largely time-invariant (e.g., sex, genetic liability, or residual confounders such as parenting practices), rather than by the reciprocal influence between constructs.

### Depressive symptoms and HDL cholesterol

With respect to lipid profiles, we detected interesting differences in the between-vs. within-person relationships between depressive symptoms and HDL cholesterol. Indeed, while the between-person correlation between random intercepts was negative (i.e., higher depressive symptoms – lower HDL cholesterol), as expected based on previous research,^44^ we found positive (reciprocal) within-person associations (i.e., higher depressive symptoms were prospectively related to higher HDL cholesterol and vice versa). While similar associations have been reported before,^45,46^ this finding was somewhat surprising and it should be interpreted with caution.

### Strengths, limitations and future directions

This is one of the very few studies capable of testing bidirectional (prospective) relationships between depressive symptoms and cardio-metabolic risk factors. To the best of our knowledge, it is the first to investigate them across such an extended developmental period. We highlighted associations with a host of cardio-metabolic risk factors (including weight- and fat-based measures of adiposity, lipid profiles and inflammatory markers), and multi-informant reports of depressive symptoms. We documented both between- and within-person relationships, which were not necessarily consistent with each other^47^ (as in the case of HDL cholesterol) and have separate implications for both mechanistic (i.e., causal) interpretations and clinical recommendations for prevention and/or intervention efforts. Finally, we provide an open-access web-application that allows researchers to explore and verify the robustness of our results.

However, these findings should be interpreted in light of some important limitations.

First, because the repeated measures leveraged by our models were collected every 1 to 6 years, these results are only informative about processes/dynamics that take place on this temporal scale. Higher temporal granularity, e.g., monthly assessments, may be needed to confirm, further refine or disprove these findings. For example, the smaller association between depressive symptoms and future adiposity, compared to that between adiposity and future depressive symptoms, could be due partly (or entirely) to inherent differences in the fluctuation of depressive mood vs. the persistence of adiposity over several years.

Second, while the inclusion of random intercepts can help reduce bias, e.g., by controlling for direct and indirect confounding effects from multiple sources,^48^ we cannot completely eliminate such bias. For example, our models do not adequately handle either non-linear nor time-varying effects of time-invariant confounders, potentially failing to account for critical biological and environmental factors at play. For example, both depressive symptoms and adiposity have been linked with endocrine processes, which are particularly salient during puberty. Future studies are therefore warranted to confirm whether these findings, are truly independent of factors such as puberty, lifestyle and/or socio-economic status.

## Conclusions

In summary, this study addresses several important limitations of previous research, which taken together may have hindered our understanding of the relationship between depressive symptoms and cardio-metabolic risk factors over the course of development. Indeed, we show that when these constructs are measured more adequately (i.e., using self-reports of depressive symptoms and fat-based measures of adiposity), their relationship appears more reciprocal than previously thought. This is especially important in light of the increasing prevalence of both early-onset depressive symptoms and child obesity, as well as their substantial, enduring consequences for life-long health and well-being.

## Data Availability

The dataset used in this study is available upon request and subject to ALSPAC executive data access procedures. To apply for access to the ALSPAC data: 1) Please read the ALSPAC access policy which describes the process of accessing the data and samples in detail, and outlines the costs associated with doing so (http://www.bristol.ac.uk/media-library/sites/alspac/documents/researchers/data-access/ALSPAC_Access_Policy.pdf); 2) You may also browse our fully searchable research proposals database, which lists all research projects that have been approved since April 2011 (https://proposals.epi.bristol.ac.uk/?q=proposalSummaries); 3) Please submit your research proposal for consideration by the ALSPAC Executive Committee (https://proposals.epi.bristol.ac.uk/).
All scripts employed in the data processing, analyses and in the generation of study metadata are publicly available (https://github.com/SereDef/comorb-longit-project).

## Acknowledgements

We are extremely grateful to all the families who took part in this study, the midwives for their help in recruiting them, and the whole ALSPAC team, which includes interviewers, computer and laboratory technicians, clerical workers, research scientists, volunteers, managers, receptionists and nurses.

## Notes

**Funding** This project received funding from the European Union’s Horizon 2020 research and innovation programme (grant reference: 848158, EarlyCause). E.W. also received funding from the UK Research and Innovation (UKRI) under the UK government’s Horizon Europe / ERC Frontier Research Guarantee [BrainHealth, grant number EP/Y015037/1] and from the National Institute of Mental Health of the National Institutes of Health (award number R01MH113930). The work of H.T. was supported by the Netherlands Organization for Health Research and Development ZonMw Vici Grant (016.VICI.170.200). The UK Medical Research Council and Wellcome (Grant ref: 217065/Z/19/Z) and the University of Bristol provide core support for ALSPAC. This publication is the work of the authors, and E.W. and S.D. will serve as guarantors for the contents of this paper. A comprehensive list of grants funding is available on the ALSPAC website (http://www.bristol.ac.uk/alspac/external/documents/grant-acknowledgements.pdf); This research was specifically funded by the Wellcome Trust and MRC (core) (Grant refs: 076467/Z/05/Z; 084632/Z/08/Z; 092731), and the John Templeton Foundation (Grant ref: 61917).

### Competing Interest Statement

The authors have declared no competing interest.

### Funding Statement

This project received funding from the European Union's Horizon 2020 research and innovation programme (grant reference: 848158, EarlyCause). E.W. also received funding from the UK Research and Innovation (UKRI) under the UK government's Horizon Europe / ERC Frontier Research Guarantee [BrainHealth, grant number EP/Y015037/1] and from the National Institute of Mental Health of the National Institutes of Health (award number R01MH113930). The work of H.T. was supported by the Netherlands Organization for Health Research and Development ZonMw Vici Grant (016.VICI.170.200). The UK Medical Research Council and Wellcome (Grant ref: 217065/Z/19/Z) and the University of Bristol provide core support for ALSPAC. This publication is the work of the authors, and E.W. and S.D. will serve as guarantors for the contents of this paper. A comprehensive list of grants funding is available on the ALSPAC website (http://www.bristol.ac.uk/alspac/external/documents/grant-acknowledgements.pdf); This research was specifically funded by the Wellcome Trust and MRC (core) (Grant refs: 076467/Z/05/Z; 084632/Z/08/Z; 092731), and the John Templeton Foundation (Grant ref: 61917).

### Author Declarations

The ALSPAC Ethics and Law Committee gave ethical approval for this study.

