## Supplementary materials for "Longitudinal co-development of mental and cardio-metabolic health from childhood to young adulthood"

### eMethods1 | Cardio-metabolic risk markers – measurement details

Besides, fat mass index (FMI), ten alternative cardio-metabolic risk markers were examined in secondary analyses. These included: lean mass index (LMI), body mass index (BMI), waist circumference, android fat mass, high-density lipoprotein (HDL) and low-density lipoprotein (LDL) cholesterol levels, triglycerides, insulin, and C-reactive protein (CRP). These were measured as follows:

Total body fat and lean mass (g) were derived from whole body dual energy X-ray absorptiometry (DXA) scans at six time points (at median ages of 9.8, 11.8, 13.8, 15.4, 17.8, and 24.5 years).[1] Android fat mass (g) was extracted from the last four DXA scans. Child weight (kg) and height (cm) were either measured during the same six research visits or reported in questionnaires (at median ages of 10.7, 12.8, 16, 17 years)[2]. Participants total body fat mass, lean mass and weight were then divided by squared height to obtain FMI, LMI and BMI respectively ($kg/m^2$). Waist circumference (cm) was measured six times (at median age of 9.8, 10.6, 11.8, 12.8, 15.4, and 24.5 years). HDL and LDL cholesterol, triglycerides, insulin and CRP were measured in non-fasting (at 9.8 years) or fasting (at 15.4, 17.8 and 24.5 years) blood samples.

**eFigure 1 | Cardio-metabolic risk markers – distributions**

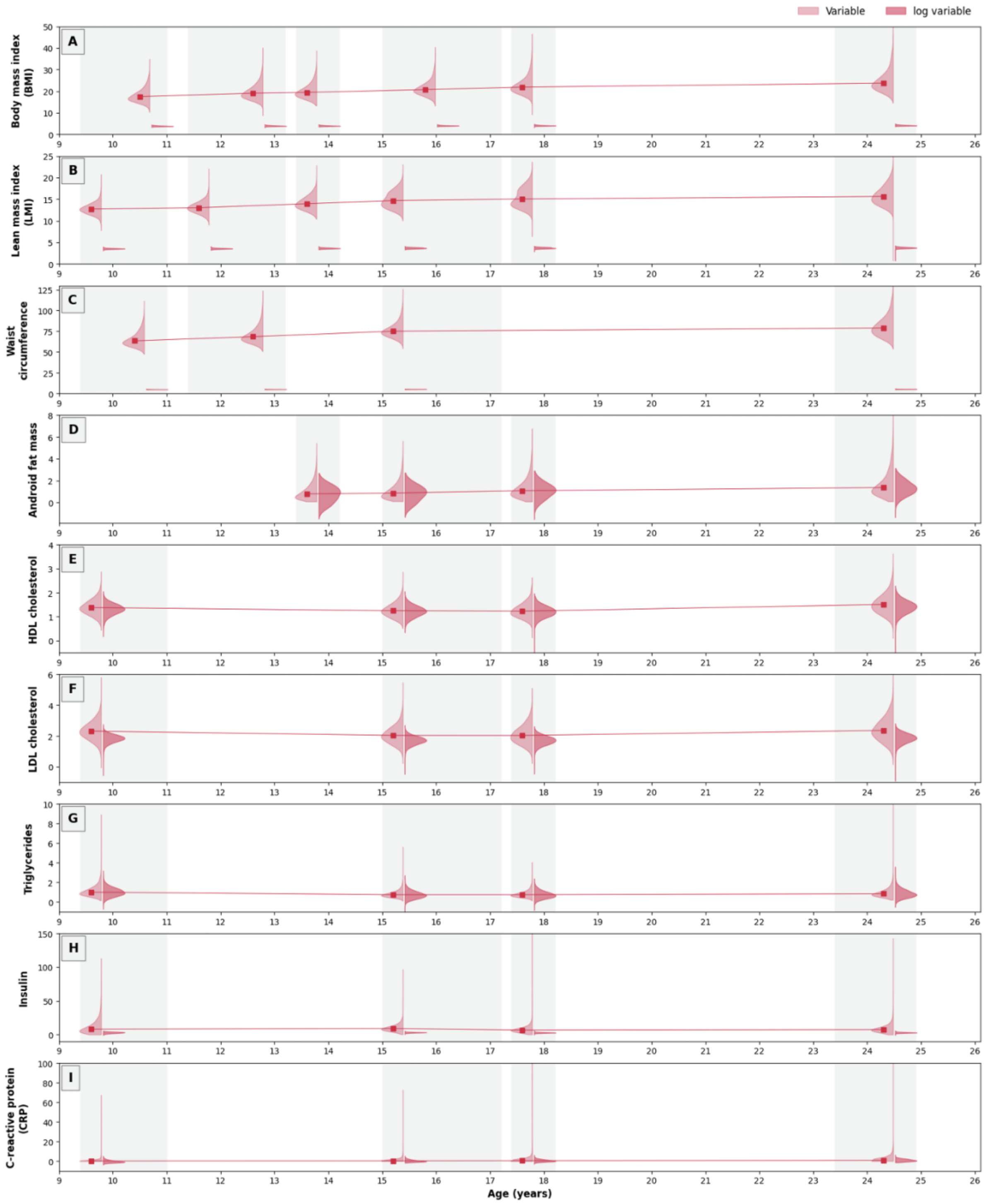

The distribution of observed values for all alternative cardio-metabolic risk markers used in secondary analyses is presented on the y-axis against measurement time (x-axis). These include (A) body mass index, (B) lean mass index, (C) waist circumference, (D) android fat mass, (E) high-density lipoprotein cholesterol, (F) low-density lipoprotein cholesterol, (G) triglycerides, (H) insulin, and (I) C-reactive protein. In the violin plots, lighter colors are used to represent the original value distributions, while darker colors represent the same variable distributions and after data transformation was applied (i.e., log transformation). The line graph connects the median points (in the original data scale) at each timepoint.

### eMethods2 | The intuition behind the (RI)-CLPM

If we conceptualize human health as a "dynamic system", which evolves over time by repeatedly driving a state (at time $t$) to another state (at time $t + 1$), we can then understand each "health state" as a function of *a)* their prior values, *b)* external inputs to the system and *c)* the transition rules or "updating mechanisms" that govern change. An example of transition rule is *persistence* (also known as inertia or self-similarity). This is the tendency for a construct to retain its state over time, until something (i.e., some external input) changes it. In traditional cross-lag panel modeling (CLPM)[3], persistence is quantified by autoregressive (AR) terms which are simply regression coefficient describing the relationship between a state and its previous values (e.g., depression at time 2 ~ depression at time 1).
Another set of transition rules, perhaps the most interesting ones from a scientific standpoint, are *reciprocal relationships*. Reciprocal relationships describe patterns of feedback across multiple variables, or the "tendency for constructs to form causal loops in which one construct initiates subsequent events that further change the original construct at a later period". For example, the onset of a depressive episode may trigger a change in body weight (e.g., because of altered inflammatory processes or changes in lifestyle) which in turn influences future states of depression (e.g., by driving self-esteem, mood or social isolation). Reciprocal relationships are also quantified in CLPMs by cross-lag (CL) regression coefficients (e.g., weight at time 2 ~ depression at time 1; *eFigure 2A*).

#### *The random-intercept CLPM (RI-CLPM): controlling for unobserved heterogeneity*

The traditional CLPM we described so far (i.e., including AR and CL terms), enjoyed long-standing popularity in psychological and epidemiological research, thanks its intuitive ability to embody core notions of system dynamics (i.e., persistence and reciprocal influence). However, this simple model has been criticised by many on account of one major methodological shortcoming: the inability to disentangle between-person effects from within-person effects, or, in other words, the lack of control for *unobserved heterogeneity*.

The random-intercept CLPM (RI-CLPM)[4] is among the more recent extensions of this framework, specifically developed to address this issue.

The RI-CLPM supposes a latent factor for each variable series (e.g., $\eta_{DEP}$ and $\eta_{CMR}$; *eFigure 2B*), i.e. the "random intercepts". These are meant to represent the collection of unmeasured confounding factors which are stable over time within people but vary from one person to the other. We refer to this concept as "unobserved heterogeneity" (but it is also known as "time-invariant confounding", "stable trait factor", or "unit fixed effects"). For example, sex, socio-economic status or genetic predisposition may be responsible for persistently higher adiposity levels across development in certain people. These stable between-person differences should not be confounded with the within-person dynamics that we mean to quantify in AR and CL terms. For this reason, the inclusion of random intercepts is a major strength of the RI-CLPM, over to the traditional CLPM, as it allows us to infer the within-person relationships between variables, that would be otherwise confounded by stable interindividual differences. The factor loadings of these time-invariant latent factors are typically set to 1 (similarly to random intercepts in fixed-effects models).

#### *The generalized CLPM (gCLPM): relaxing stationarity assumptions*

Another recent alternative to traditional CLPM, is the generalized CLPM (gCLPM) framework[5]. The gCLPM introduces two further extensions to the RI-CLPM, which provide a way to relax the assumption of stationarity, i.e., that both between- and within person effects are constant over time. In summary the model:

1) Deals with unobserved *time-varying* confounders, by allowing freely estimated $\eta$ factor loadings ("time-varying unit effects" as also mentioned in the previous paragraph). This added flexibility is important, for example, to account for major life transition periods, such as puberty, which may introduce interindividual differences (i.e., unobserved heterogeneity) that are not constant but rather have different effects across time (e.g., at age 10 vs. age 15).
2) Expands the range of temporal dynamics that the model can capture, by introducing *moving average* (MA) autoregressive and cross-lagged terms. These are time-varying coefficients, which act as direct effects of the residuals of each observed variable (known as "random impulses"). The implication of estimating these additional terms is that the influence of a predictor can be decomposed into a) a component that is stable over time, and b) a component that can differ over the various time waves. For example, adiposity may have a small influence on future depression in early adolescence but became a stronger contributor as people age.

Note that the gCLPM is not free of criticisms (see for example Usami, 2021[6]), and it is important to note how the increased flexibility may come at the cost less robust modelling results that are considerably more complex to interpret. Nonetheless, for readers who are interested in more complex characterizations of the range of dynamic processes underlying these relationships, we provide the gCLPM estimates for all variable of interests on the project web-application *[link redacted]*.

**eFigure 2 | CLPM and RI-CLPM**

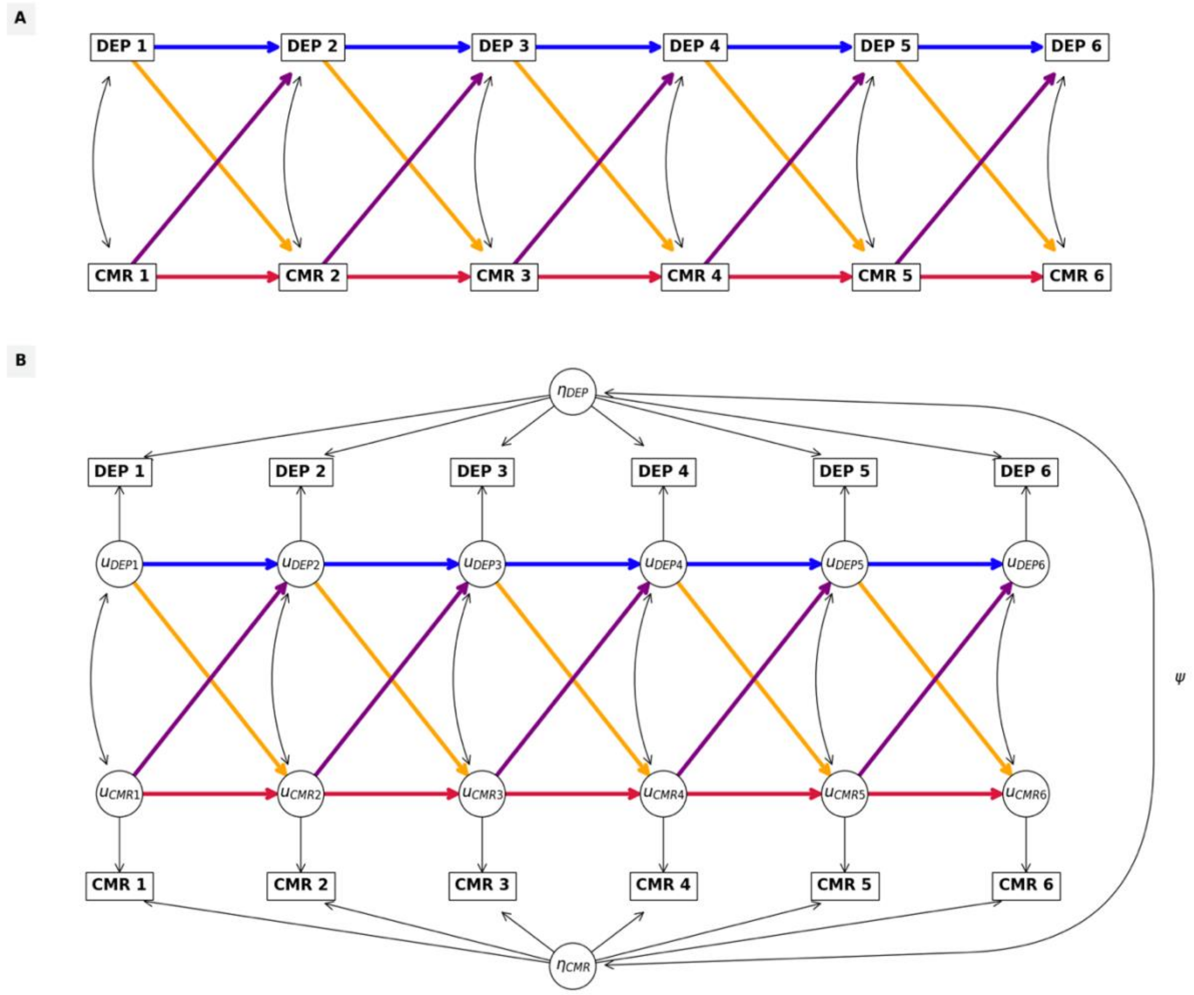

Graphic representation of (A) the traditional CLPM and (B) the RI-CLPM, described in *eMethods2*. Autoregressive (AR) terms are represented in blue and red, for depression (DEP) and cardio-metabolic risk (CMR) respectively. Cross-lag (CL) terms are depicted in yellow and purple, for depression to CMR and CMR to depression respectively.

**eFigure 3 | Cross-lag associations between maternal reports of depressive symptoms and cardio-metabolic risk factors**

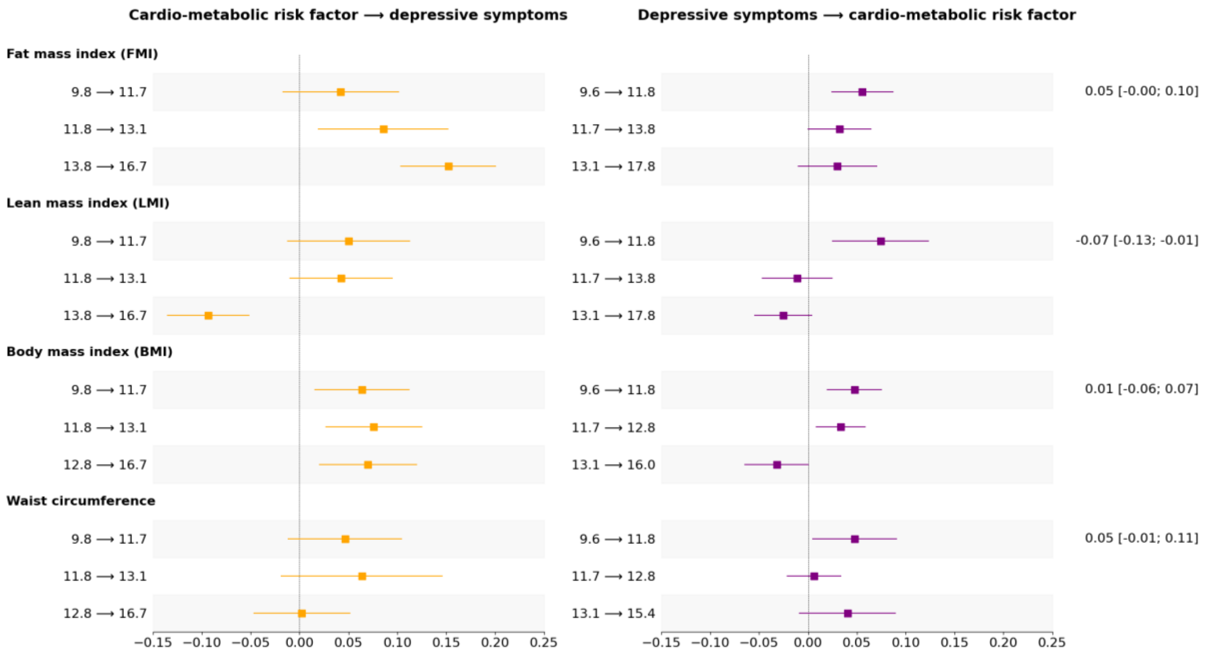

The standardised regression coefficients for the within-person cross-lag associations (and their 95% confidence intervals) are presented along the x-axis, for each cardio-metabolic risk factor examined, listed on the y-axis. The temporal lag each estimate refers to is specified in years on the y-axis. Association estimates from cardio-metabolic risk factor to lagged depressive symptoms are presented in yellow on the left and those between depressive symptoms and lagged cardio-metabolic risk factor are shown in purple on the right. In the last column on the right of the graphs, the estimated correlation coefficient between the random intercepts of each construct (and its 95% confidence interval) is reported.
